# Challenges in identifying and quantifying country-level participation in multi-country randomised controlled trials involving Ireland as a collaborating partner

**DOI:** 10.1101/2024.03.04.24303711

**Authors:** James Larkin, Uchechukwu Alanza, Vikneswaran Raj Nagarajan, Maurice Collins, Sami Termanini, Emmet Farrington, Barbara Clyne, Tom Fahey, Frank Moriarty

## Abstract

**Background and Objective:** Randomised controlled trials (RCTs) provide vital information about healthcare interventions. Accurate reporting is vital for effective RCT dissemination. This study aimed to assess the reporting quality of multi-country RCTs, using Ireland as a case study, examining trial characteristics, adherence to reporting standards and the reporting of participation from Ireland.

**Study Design and Setting:** This is a secondary analysis of RCTs identified in a previous observational study of RCTs where ≥80% of participants were recruited in Ireland. This current study focuses on multi-country RCTs with Ireland as a participating country. The current study involved an additional screening process according to these inclusion criteria: RCTs conducted on humans in a healthcare setting with centres based in Ireland, and <80% of participants recruited in Ireland. The primary outcome variables were trial characteristics and reporting rates for: trial registration, use of standardised reporting guidelines, number of Irish centres and number of participants recruited in Ireland. Descriptive statistics were used for analysis.

**Results:** Overall, 239 RCTs were included. The most common intervention was a drug (74.9% of RCTs). The most common setting was an ambulatory setting (74.1% of RCTs). The most common clinical domain was the cardiovascular system (18.0% of RCTs). Among RCTs published after the CONSORT reporting guideline was published (1996), 8.3% referred to a standardised reporting guideline. Among RCTs published after the International Committee of Medical Journal Editors mandated clinical trial registration (2005), 81.8% provided registration numbers. Number of Irish centres was reported in 75.3% (N=180) of RCTs. Number of participants recruited in Ireland was reported in 27.2% (N=65) of RCTs.

**Conclusion:** Our findings show deficits in reporting quality for multi-country RCTs, particularly in referring to reporting guidelines and reporting number of participants for the examined country. Institutions should create policies to ensure transparent RCT dissemination.

## 1. Introduction

Randomised controlled trials (RCTs) are essential for healthcare systems, in providing evidence about the safety and efficacy of interventions [1, 2]. Accurate reporting of RCTs is needed to facilitate critical appraisal, and for evidence to be acted upon. To encourage standardisation of reporting of RCTs, in 1996, the Consolidated Standards of Reporting in Trials (CONSORT) reporting guideline was published [3]. CONSORT is “a checklist of essential items that should be included in reports of RCTs,” [4] and is required by hundreds of journals [5, 6]. This includes items ranging from specifying outcomes of interest, approaches to randomisation, allocation concealment, and the setting(s) where the trial was conducted. Transparently reporting these items facilitates assessment of an RCT’s internal and external validity [5, 6]. For example, a clinician can evaluate whether the findings may be generalisable to their patient population, or whether the standard of care in the setting of the trial could impact the results. This particularly applies to multi-centre RCTs, which are increasingly ubiquitous in health research, however deficits in reporting of aspects of the multi-centre design are common [7, 8], prompting the development of multi-centre trial extensions to existing reporting guidelines [9].

Further measures that ensure transparent RCT reporting include protocol publication and pre-registering RCTs on a public trials registry. The latter, pre-specifying methods and all outcomes for an RCT and registering those details on a publicly accessible site prior to commencing patient enrolment [10], has been a publication requirement of the International Committee of Medical Journal Editors (ICMJE) since 2005, and is endorsed by various organisations including the WHO [11]. Using reporting guidelines and pre-registering RCTs aims to address several issues in RCT publications including publication bias and outcome switching [10]. However, despite these initiatives, reporting and registration issues remain [12, 13].

As well as enabling critical appraisal, clear reporting of the setting of RCTs in publications and registrations can also allow trial activity in different countries to be ascertained. This study uses Ireland as a case study to examine multi-country RCTs. RCTs are a central focus of health research in Ireland [14]. However, it is unclear whether the extent of a country’s involvement in international RCTs can be ascertained, for example, the number of participants recruited or study centres within a specific country. A 2020 study examined RCTs published before 2019 where it was reported that at least 80% of participants were recruited in Ireland [15]. Among 752 unique RCTs, it found a range of issues such as limited reference to standardised reporting guidelines, though it highlighted that the quality of reporting was improving over time [15]. That study did not examine RCTs where less than 80% of participants were recruited in Ireland.

Therefore, the aim of this study is to evaluate the reporting quality of multi-country, patient or population health-oriented RCTs. A single country is used as a case study, the study focuses on RCTs involving Ireland, examining their characteristics, their adherence to reporting standards and the level of Irish participation.

## 2. Methods

This is a secondary analysis, examining RCTs identified in a previous observational study evaluating quality and scope of RCTs in health research conducted in Ireland [15]. This previous study included RCTs where 80% or more of participants were recruited in Ireland [15]. The study drew on systematic review methods, searching six databases (e.g. PubMed), from inception to December 2018, and title/abstracts were reviewed followed by full-texts [15].

As part of the screening process for the previous observational study, RCTs were identified that had an indication of Irish participation (i.e. participants/centres/investigators from Ireland, or an author with an affiliation in Ireland) but did not clearly report at least 80% of participants recruited in Ireland. That screening process identified 428 RCTs, which were screened for inclusion in the present study based on criteria shown in table 1.

**Table 1.** Inclusion criteria for randomised controlled trials (RCTs)

| Area | Criteria |
| --- | --- |
| Population | RCTs conducted on human subjects of any age group in a health care setting (hospital/community) with at least one centre based in Ireland, and less than 80% of all participants recruited from centres based in Ireland |
| Intervention | Any intervention targeted at patients, providers and/or policy makers intended to improve health/health care (including biomedical research and population health and health services research), with no limitation on the comparator used |
| Primary outcome | Any health outcome focused on patient/population health. Studies were excluded if they only included outcomes with no direct clinical relevance (e.g. drug levels assessed in the context of a pharmacokinetic study) |
| Study design | A multi-country randomised controlled trial of any design (e.g. parallel, cluster, factorial, cross-over, stepped wedge), with two or more treatment groups |
| Publication type | Peer-reviewed research paper reporting the main results of a trial |
| Context | Conducted in two or more countries (including Ireland) |

In assessing the population, any information in the publication (including in supplementary materials) indicating participants, study centres, or investigators from Ireland were taken as meeting this eligibility criterion (unless it was explicitly stated that no participants were recruited in Ireland or all recruited from Ireland). Where a trial registry number was included, the trial registry was checked for details of any centres in Ireland. Where conflicting information was presented in the publication and the registry, information from the publication was used. Publications were excluded where the only indication of participation from Ireland was an author with an affiliation at an Irish institution, but no further information that indicated participants recruited in Ireland. If the publication found was a study protocol, or interim, secondary, or sub-group analysis, a supplementary search was conducted (details in Appendix A eBox 1) to identify the main publication.

### 2.1 Study inclusion and data extraction

Titles and abstracts for the 428 RCTs identified by Clyne and colleagues [15] were reviewed independently by two reviewers (EF or FM) who excluded any studies that clearly did not meet inclusion criteria. Full-texts for remaining publications were reviewed for eligibility by one reviewer (MC, VRN, UA, ST) according to the inclusion criteria. Data relating to participation from Ireland was checked by a second author (JL) in all cases. Any instances where it was unclear whether there was Irish participation were discussed with FM or TF.

A data extraction form, in Microsoft Excel, was piloted by two reviewers (FM and TF) and modified before the main data extraction phase. The extraction form included the following variables: publication information, eligibility, study documentation (e.g. ethics approval), study characteristics, methods, participants, and results. A standardised operating procedure for data extraction (details in Appendix A, eTable 1) was developed and read by reviewers before commencing, and this was iteratively updated with further guidance as queries arose. Data was extracted by one reviewer (MC, VRN, UA, ST, or JL), with 10% also reviewed and extracted by a second reviewer (FM) to assess consistency.

### 2.2 Variables

There were several variables extracted, across the following categories: 1) Study characteristics and methods, 2) Study documentation and reporting, and 3) Countries, centres and participants (see Appendix A, eTable 2).

#### 2.2.1 Study characteristics and methods

Data on study design, unit of randomisation, setting, population, intervention and comparator type, funding source, and primary outcome. Primary outcome was categorised using the core areas within the outcome taxonomy developed by Dodd and colleagues: 1) physiological/clinical, 2) life impact, 3) death, 4) adverse events 5) resource use [16] and a sixth category that we added: other. Clinical domain was categorised using the UK Clinical Research Collaboration’s Health Research Classification System [17]. Other variables included, whether baseline characteristics were presented and whether the primary outcome result was positive or negative.

#### 2.2.2 Study documentation and reporting

The study documentation variables included whether there was reference to 1) ethical approval, 2) trial registration number, 3) a standardised reporting guideline, 4) publication of protocol, and 5) a sample size calculation (Appendix A, eTable 2).

#### 2.2.3 Countries, centres and participants

The main outcomes for Irish participation in the included RCTs were assessed as quantification of reporting of each of the following:

- Centres in Ireland among the total number of included centres
- Participants recruited from Ireland among the total number of participants

### 2.3 Analysis

The study characteristics and methods of included RCTs were summarised using frequencies and percentages. Also, median (interquartile range) number of participants recruited overall and in Ireland for each characteristic was calculated. Study documentation and reporting was summarised as the number and percentage of RCTs referring to ethics approval, a standardised reporting guideline, published protocol or trial registration number, and where applicable, restricting to years since this provision was introduced. Percentage of RCTs with each was also plotted over time. For countries, centres, and participants, the number and percentage of RCTs reporting Irish participant at each level was summarised. The median (IQR) number and percentage of Irish centres and participants were also calculated, and violin plots generated to visualise the distribution of these. For countries, a network analysis was conducted to examine the pattern of countries participating in RCTs with Ireland. Lastly, the level of Irish participation reported in RCTs was summarised based on presence of key study documentation and funding source.

## 3. Results

From the 428 RCTs reviewed, 168 were excluded (47 at title/abstract screening, and 134 at full-text review), most often as secondary analyses, duplicates or not reporting participants recruited in Ireland (see Figure 1). A further 21 RCTs that had a co-author based in an Irish institution but no other indication of Irish involvement were excluded.

**Figure 1.**
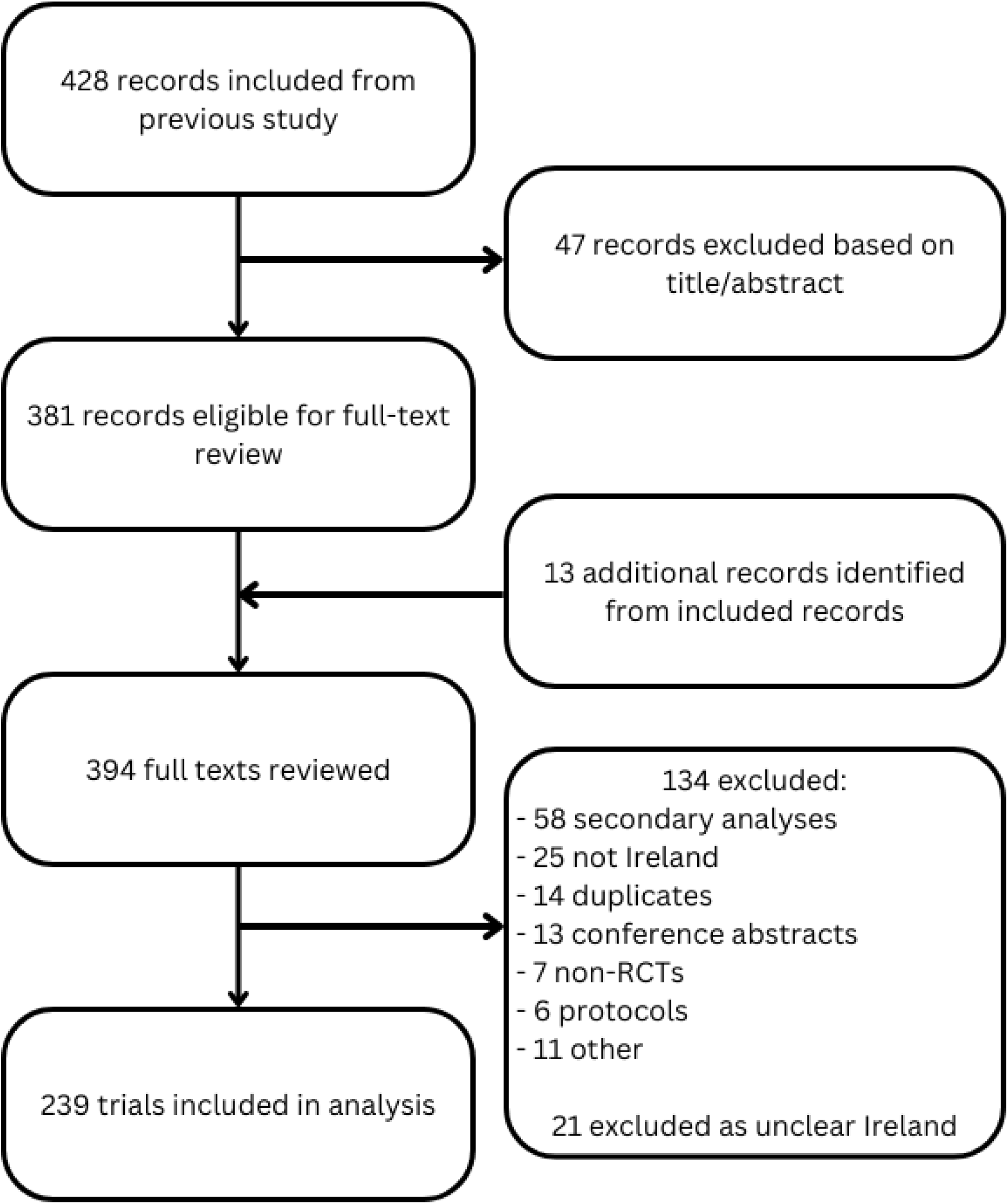
Illustration of inclusion of randomised controlled trials in this study.

Overall, 239 studies met the inclusion criteria. For two included studies, the primary publication could not be retrieved and therefore associated publications (e.g. secondary analysis) were used to extract relevant information. However, it was therefore not possible to make conclusions on several variables for these RCTs (details in appendix A, eTable 3).

### 3.1 Study characteristics and methods

RCTs were mainly parallel design (92.1%, N=220), evaluated drugs (74.9 %, N=179) among adults (74.9%, N=179) in an ambulatory (outpatient) setting (74.1%, N=177) (Table 2). The most common funding source was industry (54.8%, N=131), randomisation primarily occurred at the individual level (96.2%, N=230), the most common type of blinding was double-blinding (49.8%, N=119), and the primary outcome measure in 56.1% (N=134) of RCTs was categorised as physiological/clinical.

**Table 2.** Study characteristics.

| Characteristics | Overall % (N, RCTs) | Number of participants overall Median (IQR) | Number of participants recruited in Ireland Median (IQR) |
| --- | --- | --- | --- |
| **RCT Design** |  |  |  |
| Parallel | 92.1% (N=220) | 412 (224-960) | 49 (12-74) |
| Factorial | 4.2% (N=10) | 892 (456-1,367) | 80 (72-88) |
| Crossover | 3.3% (N=8) | 78 (47-290) | N.A. |
| Stepped Wedge | 0.4% (N=1) | N.A. | N.A. |
| **Clinical Setting** |  |  |  |
| Ambulatory (Outpatient) | 74.1% (N=177) | 409 (249-940) | 39 (11-64) |
| Hospital (In-patient) | 17.6% (N=42) | 330 (155-927) | 34 (12-68) |
| Primary care | 4.2% (N=10) | 784 (464-2,077) | 68 (36-99) |
| Multiple sites | 2.9% (N=7) | 1041 (528-7,201) | 79 (70-95) |
| Other | 1.3% (N=3) | 124 (102-930) | 50 (50-92) |
| **Population** |  |  |  |
| Adults | 74.9% (N=179) | 417 (223-973) | 53 (24-79) |
| Older adults | 8.8% (N=21) | 433 (351-1,759) | 29 (10-182) |
| Children | 5.4% (N=13) | 262 (216-521) | 7 (5-10) |
| Infants | 5.4% (N=13) | 166 (108-570) | 14 (4-26) |
| Pregnant people | 4.2% (N=10) | 877 (413-14,397) | 8 (4-449) |
| Multiple populations | 1.3% (N=3) | 618 (392-824) | 1 (1-1) |
| **Intervention** |  |  |  |
| Drug | 74.9% (N=179) | 417 (227-1,008) | 28 (11-68) |
| Procedure/Surgery | 9.2% (N=22) | 340 (140-896) | 17 (4-48) |
| Supplement | 4.2% (N=10) | 352 (288-433) | 64 (59-147) |
| Behavioural | 3.8% (N=9) | 529 (436-1,736) | 100 (75-117) |
| Health System Intervention | 3.3% (N=8) | 661 (320-1,394) | 79 (61-131) |
| Device | 1.3% (N=3) | 108 (93-115) | 29 (16-41) |
| Biological/Vaccine | 1.3% (N=3) | 349 (332-1,736) | N.A. |
| Mixed | 2.1% (N=5) | 515 (324-1,010) | N.A. |
| **Funding source** |  |  |  |
| Industry | 54.8% (N=131) | 391 (238-855) | 38 (16-74) |
| Government body/agency | 17.6% (N=42) | 618 (324-1,549) | 42 (5-64) |
| Not stated | 13.8% (N=33) | 208 (84-469) | 49 (26-50) |
| Multiple funders | 8.8% (N=21) | 2,004 (666-3,222) | 57 (29-131) |
| Charity | 2.1% (N=5) | 274 (262-333) | 56 (34-78) |
| University | 0.8% (N=2) | 1,070 (605-1,534) | N.A. |
| Explicitly report no funding | 0.4% (N=1) | N.A. | N.A. |
| Other/Unsure | 1.7% (N=4) | 78 (67-415) | N.A. |
| **Randomisation level** |  |  |  |
| Individual | 96.2% (N=230) | 412 (222-986) | 49 (12-74) |
| Group/cluster | 3.8% (N=9) | 629 (361-2,454) | 75 (46-103) |
| **Blinding** |  |  |  |
| Double-blinding | 49.8% (N=119) | 380 (208-944) | 34 (15-87) |
| Single-blinding | 11.7% (N=28) | 330 (163-536) | 60 (34-64) |
| Open label | 38.1% (N=91) | 452 (260-1,021) | 49 (11-82) |
| Other | 0.4% (N=1) | N.A. | N.A. |
| **Primary Outcome Category** |  |  |  |
| Physiological/clinical | 56.1% (N=134) | 346 (204-683) | 51 (8-79) |
| Life impact | 5.0% (N=12) | 466 (124-1,159) | 52 (42-73) |
| Death | 4.6% (N=11) | 1,363 (676-4,446) | 82 (44-164) |
| Adverse events | 0.4% (N=1) | N.A. | N.A. |
| Mixed | 31.4% (N=75) | 510 (310-1,351) | 29 (12-63) |
| Other | 2.5% (N=6) | 560 (77-3,238) | 38 (21-52) |

Abbreviations: N.A.=Not applicable

The baseline characteristics of participants were presented in 90.0% (N=215) of RCTs. In 25.1% (N=60) of RCTs the comparison group used was placebo drug or sham intervention (full details in Appendix A, eTable 4). The result for the primary outcome was positive in 54.0% (N=129) of RCTs, negative in 34.3% (N=82) and for 11.7% (N=28) of RCTs insufficient information was provided to make a conclusion. The cardiovascular system was the focus of 18.0% (N=43) of RCTs (Figure 2).

**Figure 2.**
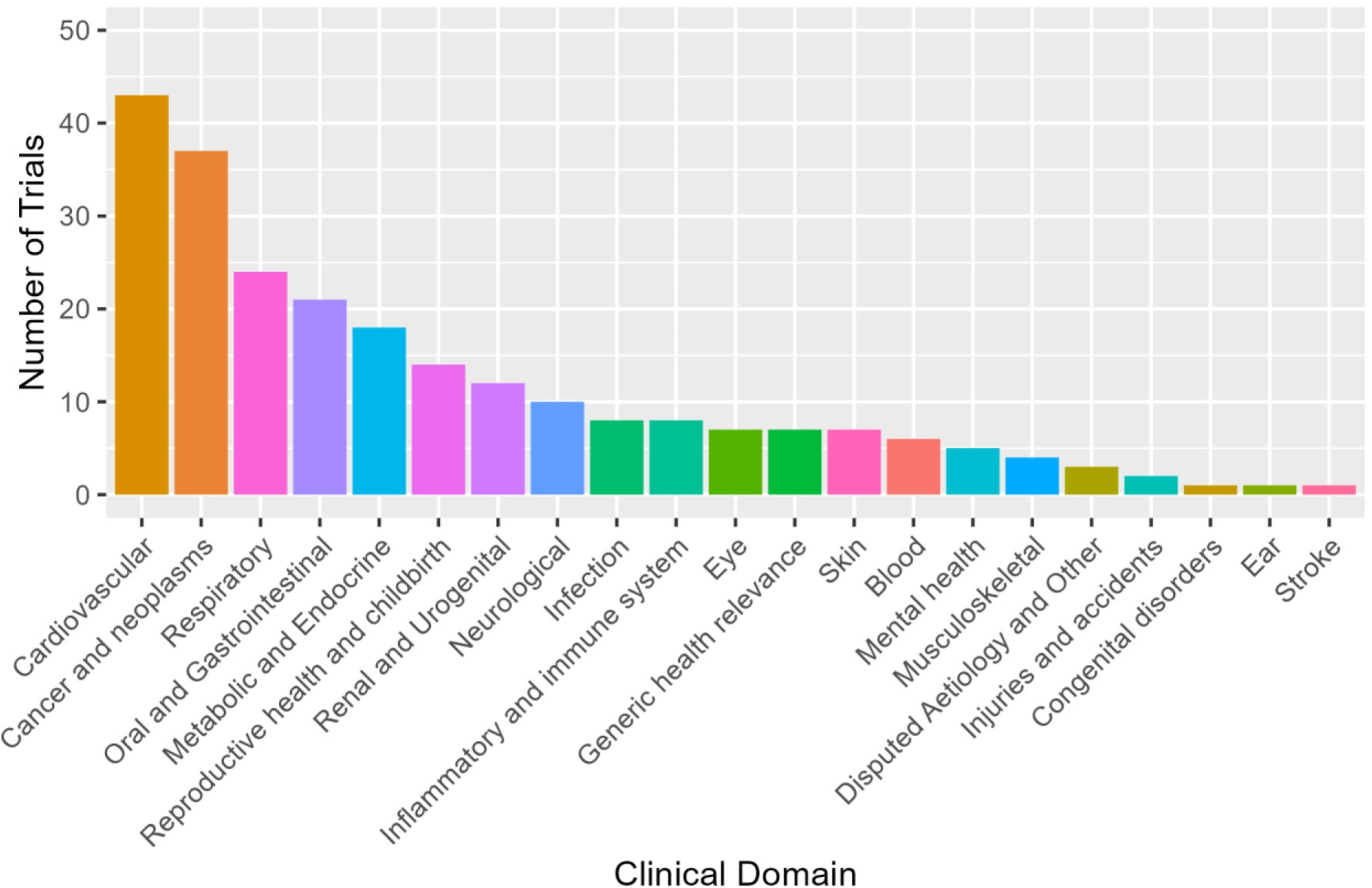
Clinical domains of included randomised controlled trials classified using UK Clinical Research Collaboration’s Health Research Classification System

### 3.2 Study documentation and standardised reporting

A published protocol was referred to in 27.3% (N=65) of RCTs. Of the 204 RCTs published after 1996, when the CONSORT statement was published, 8.3% (N=17) referred to a standardised reporting guideline. Of the 132 RCTs that were published after 2005, when the ICMJE mandated clinical trial registration, 81.8% (N=108) provided a trial registration number. It was stated that the trial had ethical approval in 92.9% (N=221) of RCTs. Figure 3 provides detail of reference to trial registration, protocol publication, reporting guidelines, and ethical approval, across years.

**Figure 3.**
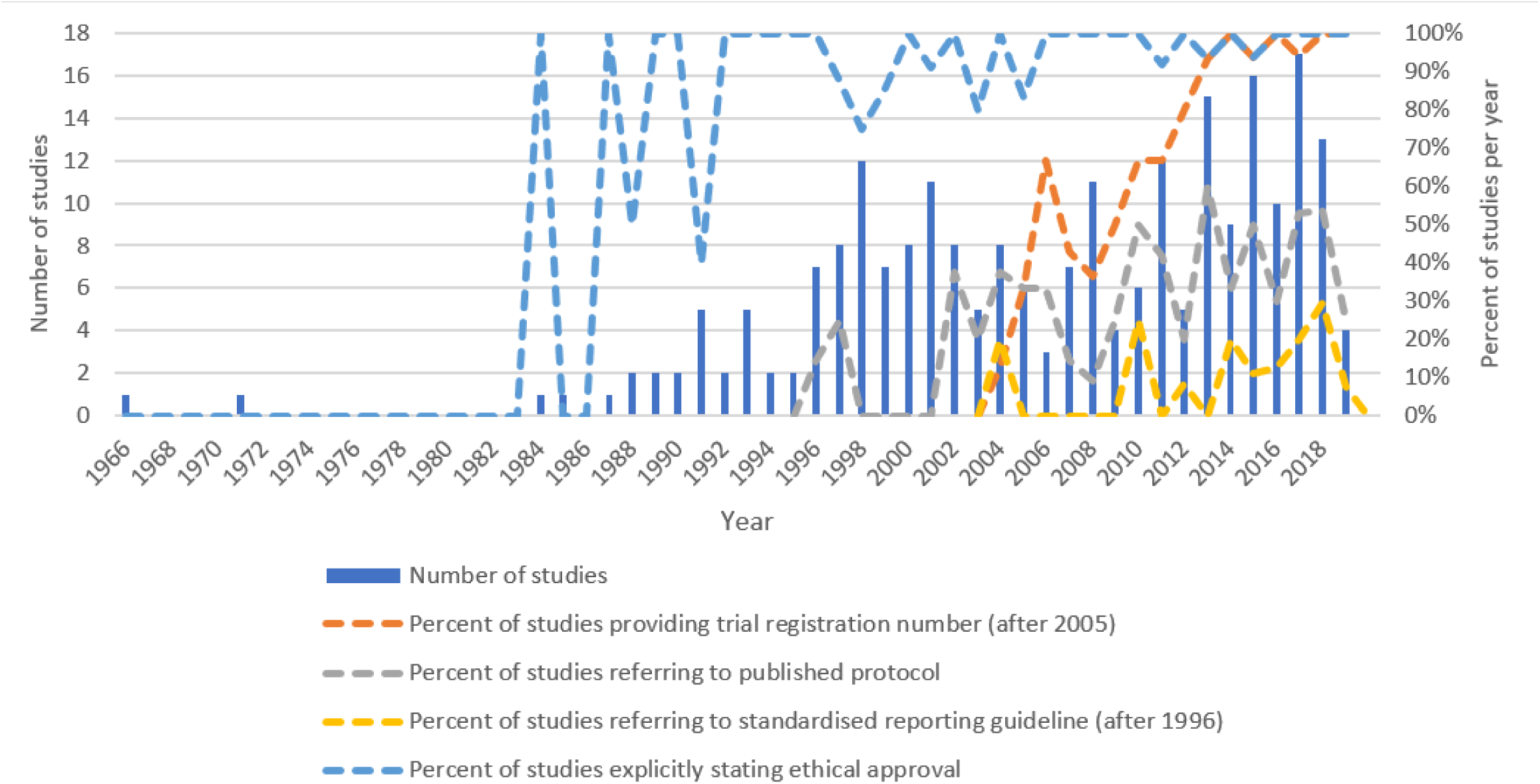
Number of studies published and percentage meeting reporting standards per year For 67.4% (N=161) of RCTs, information about a sample size calculation was provided, and amongst these, the required sample size was reached in 76.4% (N=123) of RCTs. The required sample size was not reached in 20.5% (N=33) of RCTs, and for 3.1% (N=5) of RCTs there was reference to a sample size calculation but an exact figure was not provided.

### 3.3 Countries, centres and participants

Information on the number of countries that participants were recruited from was provided in 100% (N=239) of RCTs. Source of information on countries is in Appendix A, eBox 1. Across included RCTs, there were 90 different countries (excluding Ireland) that recruited participants (full details in Appendix A, eTable 5). The median number of countries other than Ireland was 4 (IQR:1-11). Ireland made up a median of 20% (IQR:8.3%-50%) of countries involved in RCTs. The United Kingdom was the country that was included in the most RCTs (81.2%, N=194), followed by Germany (39.3%, N=94). Figure 4 illustrates participation between the top 20 countries.

**Figure 4.**
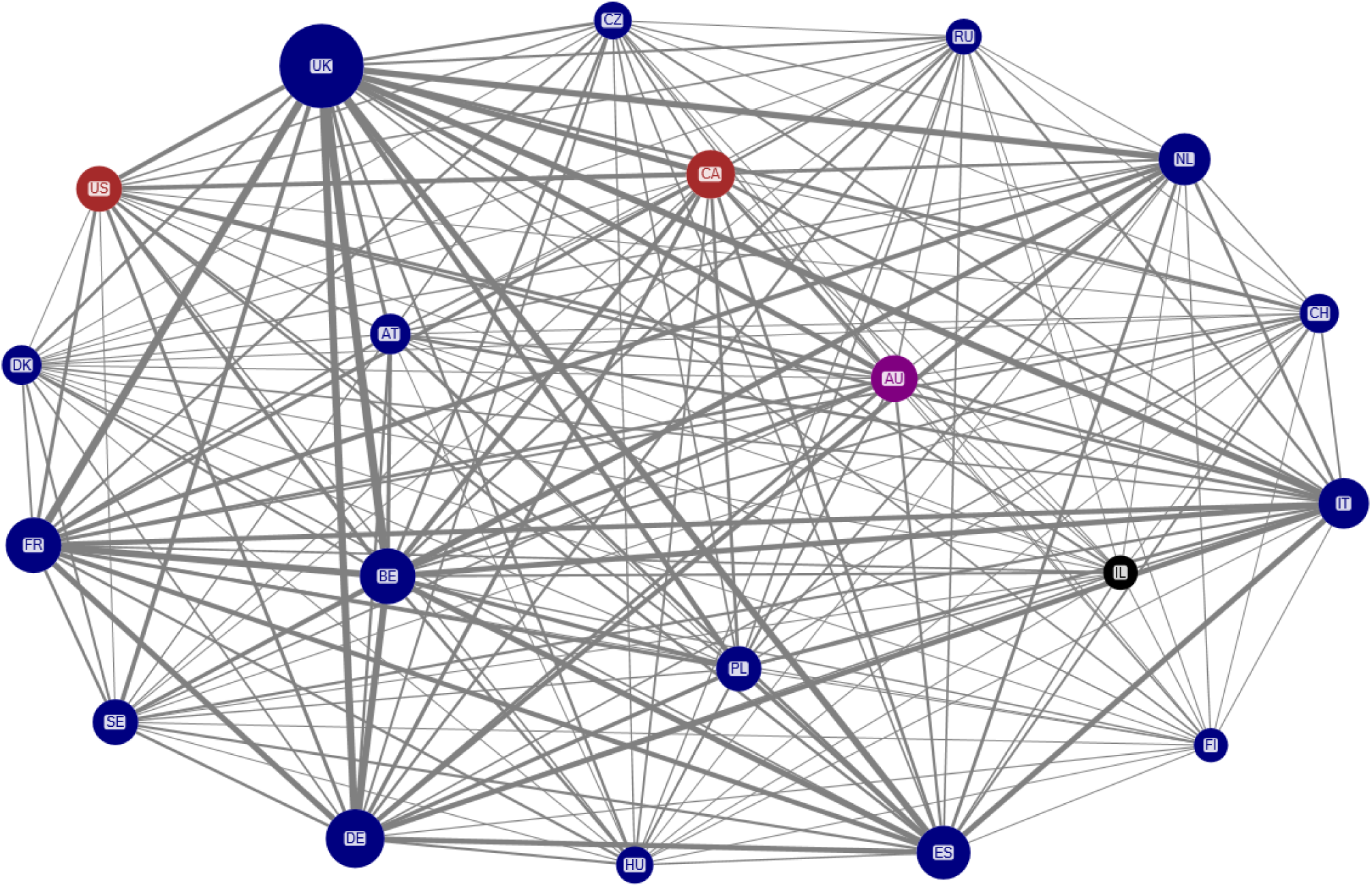
Network analysis of the 20 countries that appeared most frequently in randomised controlled trials Note: Node size is proportional to the number of randomised controlled trials, line thickness is proportional to the number of collaborations. Node colours: North America = red; Europe = navy; Oceania = purple; Asia = black. Countries are named using two letter country codes.

Information was provided on the total number of centres in 92.1% (N=220) of RCTs. The median number of centres was 33 (IQR:13-101). The number of Irish centres was reported in 75.4% (N=180) of RCTs. The median number of Irish centres was 1 (IQR:1-3). For RCTs that reported both the overall number of centres and the number of Irish centres, 74.9% (N=179) of RCTs, the median percentage of centres in Ireland was 6.0% (IQR:2.2%-15.4%). RCTs reported number of Irish centres (75.4%, N=180) more often than they reported list of investigators involved in the trial (68.2%, N=163).

All included RCTs provided information on the number of participants randomised; the median was 417 (IQR:223-1,004). Information was provided on the number of participants recruited in Ireland in 27.2% (N=65) of RCTs. The levels of reporting for countries, centres and participants is summarised in Figure 5. The median number of participants recruited in Ireland was 49 (IQR:12-79). The median percentage of participants that were recruited in Ireland was 5.3% (IQR:1.7%-17.1%). The distribution of the proportion of Irish centres and participants recruited in Ireland is in Appendix A, eFigure 1.

**Figure 5.**
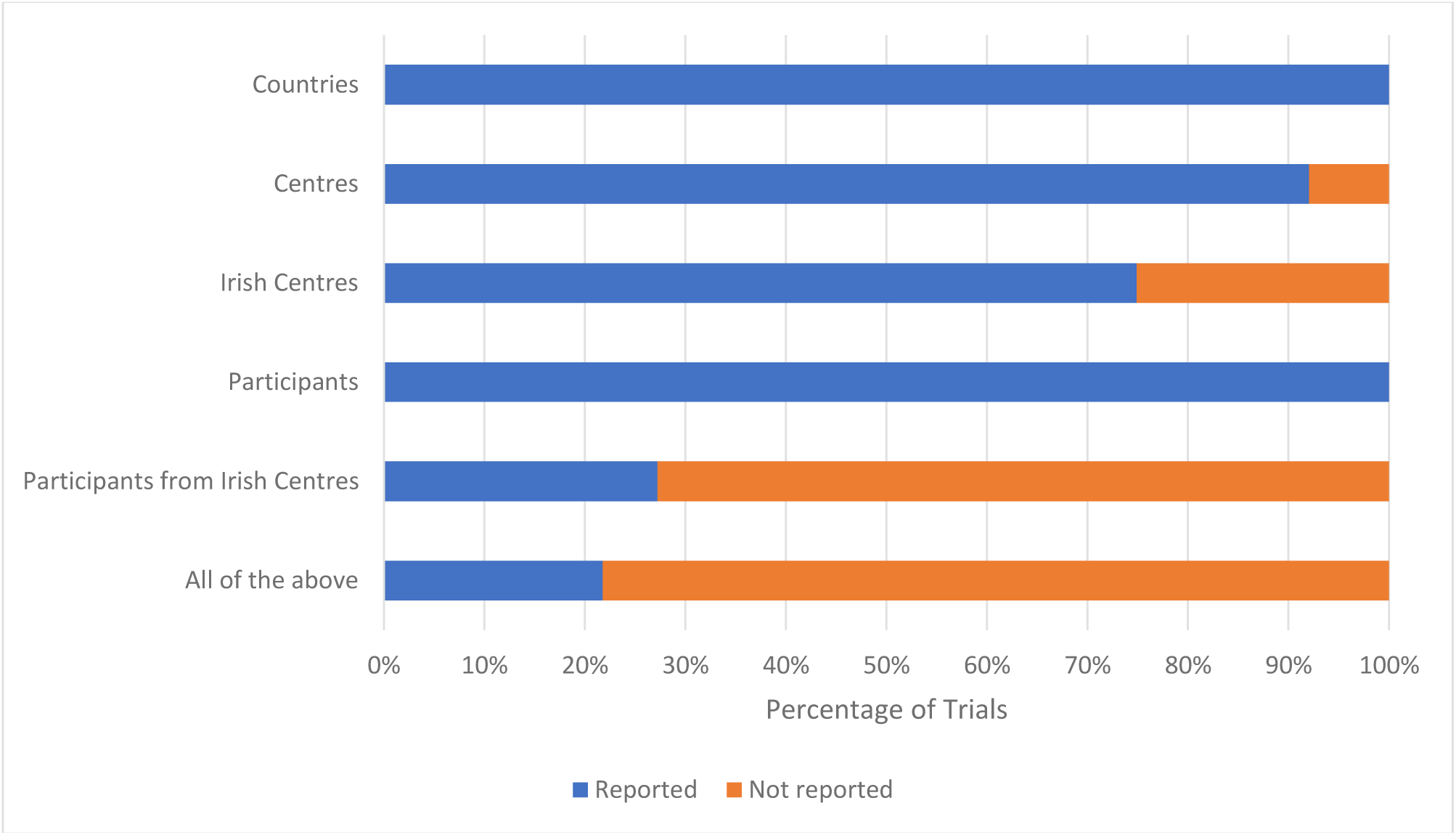
Reporting levels for centres, participants and countries.

Studies that provided details of ethical approval, trial registration number, reporting guideline used or a published protocol had higher percentages of reporting number of Irish centres and number of participants recruited in Ireland (Table 4).

**Table 4.** Level of reporting of participants recruited in Ireland and Irish centres across trial characteristics.

|  | Reporting number of participants recruited in Ireland<br>N (%) | % of participants recruited in Ireland<br>Median (IQR) | Reporting number of Irish centres<br>N (%) | % of Irish centres<br>Median (IQR) |
| --- | --- | --- | --- | --- |
| Reporting guideline referenced (1997 onwards) | 58.5% (N=10) | 6.2% (2.7%-7.6%) | 82.4% (N=14) | 9.7% (4.3%-13.8%) |
| Reporting guideline not referenced (1997 onwards) | 25.7% (N=48) | 4.6% (1.7%-15.9%) | 75.9% (N=142) | 4.9% (2.0%-13.3%) |
| Ethical Approval mentioned | 28.5% (N=63) | 5.2% (1.6%-12.9%) | 76.5% (N=169) | 5.8% (2.2%-15.0%) |
| Ethical Approval not mentioned | 11.8% (N=2) | 30% (24.9%-35.2%) | 58.8% (N=10) | 8.5% (2.4%-23.8%) |
| Trial registration number provided (2006 onwards) | 35.2% (N=38) | 4.4% (1.5%-7.9%) | 88.9% (N=96) | 3.8% (2.0%-10.2%) |
| Trial registration number not provided (2006 onwards) | 33.3% (N=8) | 25.3% (19.4%-41.1%) | 62.% (N=15) | 7.7% (2.9%-4.2%) |
| Published protocol mentioned | 43.1% (N=28) | 2.5% (1.2%-11.8%) | 84.6% (N=55) | 3.5% (1.7%-8.9%) |
| Published protocol not mentioned | 21.4% (N=37) | 5.8% (3.7%-18.5%) | 71.7% (N=124) | 7.7% (2.8%-20.0%) |
| **Funding source** |  |  |  |  |
| Industry | 20.6% (N=27) | 4.6% (2.0%-16.0%) | 76.3% (N=100) | 4.7% (1.9%-9.6%) |
| Government body/agency | 52.4% (N=22) | 4.7% (1.3%-7.6%) | 71.4% (N=30) | 8.9% (4.3%-9.8%) |
| Not stated | 15.2% (N=5) | 40.2% (34.3%-40.4%) | 66.7% (N=22) | 21.0% (5.2%-33.3%) |
| Multiple funders | 42.9% (N=9) | 2.4% (1.4%-5.3%) | 85.7% (N=18) | 4.5% (1.7%-10.3%) |
| Charity | 40.0% (N=2) | 17.3% (10.9%-23.7%) | 80.0% (N=4) | 3.5% (3.4%-15.2%) |
| University | 0.0% (N=0) | N.A. | 100.0% (N=2) | 12.0% (6.9%-17.1%) |
| Explicitly report no funding | 0.0% (N=0) | N.A. | 0.0% (N=0) | N.A. |
| Other /Unsure | 0.0% (N=0) | N.A. | 75.0% (N=3) | 25.0% (17.3%-37.5%) |

## 4. Discussion

This observational study of multi-country RCTs, focusing on those involving Ireland as a case study, identified 239 unique RCTs published between 1966-2019. Our findings suggest that both the number of multi-country RCTs and the reporting quality of multi-country RCT articles have increased steadily over time. However, deficits in reporting quality remain. Using our case study, Ireland, as an example, we found that reporting to quantify country involvement was very limited; only 27% of RCTs provided the number of participants recruited in Ireland and the number of Irish centres was reported in 75% of RCTs. Information on the total number of centres was provided in 92% of RCTs, comparable to an analysis of 82 RCTs published in five major journals in 2015 which found that 94% of RCTs stated the number of centres, suggesting the sample of RCTs in the present study is comparable to the wider literature [18]. Overall, this research is part of a growing focus on regional/country analyses of trial conduct and reporting [19, 20]. There were other important reporting issues: only 8% of RCTs published after 1996 (when the CONSORT reporting guideline was published) explicitly referred to a standardised reporting guideline, which is low given that most major journals require adherence to such guidelines. This is not improving over time; only 8% of the 13 RCTs published in 2018 referred to a reporting guideline. This compares unfavourably to the previous Clyne study of RCTs where 80% or more of participants were recruited from Irish centres (mainly RCTs led by investigators based in Ireland), in which 16% of RCTs referenced a reporting guideline [15].

Trial registration levels in the current study are relatively high; 82% of RCTs published after 2005 referred to a trial registry number. Also, trial registration improved over time, with 100% trial registration in several recent years. This compares well to the Clyne study, which found that registration only occurred for 32% of included RCTs [15]. A 2021 survey of 2,844 multicentre RCTs, found that that the registration number was provide in <50% of RCTs [8]. The Irish investigator-led RCTs compare poorly with regard to published protocols too, with only 10% of RCTs referring to a published protocol [15] compared to 27% in the current study. The 2021 survey of multicentre RCTs found that details of a full protocol were available in <50% of RCTs [8].

The RCTs examined in this study took place across 91 countries. A previous study identified 154 countries contributing to almost 40,000 RCTs, and the countries contributing most were similar to those found in our study of Irish RCTs (e.g. UK and Germany) [21]. In relation to funding, industry funded 55% of the included RCTs. A study of the most cited RCTs in the years 2019-2022 found that the majority of included RCTs were funded by industry [22]. This is concerning, as industry funded RCTs are more likely to report results and conclusions that favour the study’s intervention [23, 24].

### 4.1 Recommendations and Future Research

Funders, institutions, regulators, industry and journal editors should ensure adequate governance measures are in place to ensure transparent reporting of RCTs. This could include institutions creating a policy of mandated results dissemination, and structure to support this, as well as investing in transparency guidance and training [25, 26]. Journals should consider interventions shown to be effective for improving reporting quality such as conducting an additional peer review where the focus is on reporting quality [27, 28]. Another initiative with potential to address issues outlined in this study is the development of a CONSORT Extension for multicentre RCTs [9] which will aid reporting of multicentre RCTs.

### 4.2 Strengths and limitations

This study’s primary strength is that, along with the Clyne study [15], it provides a comprehensive overview of RCTs with centres based in Ireland. It is novel in being one of the first studies to assess the extent to which participation of centres/individuals from specific countries is quantified in trial publications. The primary limitation is that some relevant studies may not have been identified. However, many of these are likely due to poor reporting. Another limitation is that the years used to assess trial preregistration and standardised reporting may be overly strict, however we also present yearly rates (Figure 2). Also, the use of a reporting guideline, publication of a protocol or registration do not necessarily mean that these documents were completed comprehensively or adhered to [5]. However, the availability of these documents does facilitate the appraisal of the trial, though this is beyond the scope of our study. Also, some studies may adhere to a reporting guideline without explicitly referring to one. Other relevant limitations are outlined by Clyne and colleagues [15].

### 4.3 Conclusion

This study of multi-country RCTs adds further evidence that reporting and registration of RCTs is sub-optimal, though it is improving. Quantifying the participation of countries in RCTs at centre or participant-level is limited by poor reporting in multi-country RCT publications. Further improvements could be potentially made through interventions by institutions, funders and journals.

## Authors’ contributions

All authors screened studies and conducted data extraction. FM & JL prepared the database for analysis. FM & JL prepared the manuscript. TF & FM conceived the original idea for the study, and all authors reviewed and approved the final version of this manuscript.

## Data availability

The relevant data set is available at https://doi.org/10.5281/zenodo.10653711.

## Acknowledgements

We are thankful to Dr. Alan Barry for his assistance with record management.

