## Appendix A for "Challenges in identifying and quantifying country-level participation in multi-country randomised controlled trials involving Ireland as a collaborating partner"

**APPENDICES FOR:**

**Reporting standards and scope of randomised controlled trials with Ireland as the minority collaborating partner: an observational study**

By: James Larkin, Uchechukwu Alanza, Vikneswaran Raj Nagarajan, Maurice Collins, Sami Termanini, Emmet Farrington, Barbara Clyne, Tom Fahey, Frank Moriarty

eBox 1. Supplemental search strategy

In cases where the publication was a conference abstract, study protocol, or interim, secondary, or sub-group analysis, one study author (FM) conducted a supplementary search was conducted (full details in Appendix A eBox 1) in PubMed and clinicaltrials.gov using key trial details (study name/acronym, population, intervention, clinical trial registration number), to identify the main trial publication for inclusion. Searches for main study publications were conducted up to May 2023. If the full text of the main publication could not be retrieved then any available associated publications (e.g. secondary analyses or protocols) were used for data extraction.

eTable 1. Standardised operating procedure for data extraction

| **General details** | **Format** | **Instructions** |
| --- | --- | --- |
| **Data extraction by** | Prepopulated |  |
| **Study Title** | Prepopulated |  |
| **Author** | Prepopulated |  |
| **Year of publication** | Prepopulated |  |
| **Journal** | Prepopulated |  |
| **Meets inclusion criteria** | Dropdown: Yes, No, Unsure |  |
| **If doesn't meet inclusion, reason for exclusion** | Dropdown based on inclusion criteria:   - Not ROI - Unclear ROI - Conference abstract only - Study design not RCT - Duplicate - Secondary data analysis/related trial publication e.g. cost-effectiveness - Non health outcome - Protocol - Not Human - Article retracted - Other | Eligibility criteria:   1. Population: all studies conducted on human subjects of all age groups in a health care setting (hospital/ community) with at least one centre based in Ireland, and less than 80% overall recruited from Ireland. 2. Intervention: Any intervention targeted at patients and/or providers and/or policy makers intended to improve health/health care (including biomedical research and population health and health services research) 3. Comparators: no limitation on comparators 4. Primary outcome: any health outcome focused on patient/population health 5. Study design: Described by authors as a randomised controlled trials (RCT) of any design (e.g. parallel, cluster, factorial, cross-over, stepped wedge), with two or more groups.  However, author-described RCTs which did not state explicitly that random allocation was employed were managed by consensus and studies that stated information to the contrary were excluded e.g. authors state paper as RCT but describe recruitment as consecutive. Clinical trial phases II and above, so long as they were randomised. 6. The main publication relating to a trial. Some of the records identified may just be conference abstracts, protocols for studies (i.e. the study plan but no results), or secondary analyses (like economic evaluations, or further analysis of a previously published trial).   For **population from Ireland**:   - Any information in the paper indicating participants, study centres, or investigators from Ireland can be taken as meeting this eligibility criterion (unless it explicitly states no one was recruited from Ireland or all recruited from Ireland). - For all studies with a trials registry number (e.g. NCT, ISRCTN, EUCTR), please check the registry to check if Irish centres are reported. Where conflicting information is present in the paper versus the registry, please use the information from the paper. - Information on centres etc may be contained in an appendix which you may have to access on the journal website. If you can’t access this directly, try search the journal and use off campus access through RCSI library. If this does not work, email Frank with details and he will try to source this. - Where the only indication of participants from Ireland is an author with an Irish affiliation, and no registry entry, we will contact the corresponding author at a later stage.   The outcome eligibility criteria (#4) may be a little bit difficult to judge. Essentially we are looking for some sort of health outcomes, i.e. something that is useful clinically. So we would be ruling out pharmacokinetic studies as an example if they are only measuring drug levels, or more lab-based studies e.g. biomechanical studies looking at range of movement of a joint based on different exercises. If unsure whether a clinically-relevant outcome is included, mark as unsure and a second reviewer can check this.  For main publication relating to trial (#6), if the paper seems to be some form of duplicate publication  (e.g. reporting longer follow up of previously published trials, analysis of secondary  outcomes, or sub-group analyses, or conference abstracts/ study protocols), please:   - Send these on to Frank, along with details of any original trial publication cited in the paper (e.g. they may refer to the main results of the trials having been published previously and provide a reference). - Frank will try search for an original trial publication (if you did not find details of this), and will confirm whether or not this is already included in our identified papers. - If it has not already been included, Frank will let you know, and ask you to add this to your data extraction sheet and proceed with assessing eligibility and extraction. |
|  | Free text | If selected ‘Other’ above, or if unsure about exclusion, enter more details here on potential reasons for exclusion. |
| **Trial abbreviation** | Free text | Details of trial abbreviation/acronym (if applicable) |
| **Study documentation** |  |  |
| **Trial registration number** | Free text | Details of any/all trial registration numbers provided e.g. NCT, EUCTR, ISRCTN  NCT numbers can be searched for in [www.clinicaltrials.gov](http://www.clinicaltrials.gov/), EUCTR numbers in <https://www.clinicaltrialsregister.eu/>, ISCRTN numbers in <https://www.isrctn.com/>. For other registration number formats, a Google search will identify the relevant registry. |
| **Refers to published protocol** | Dropdown: Yes, No, Unsure | Does the paper either provide a link to a protocol or reference a protocol published in a peer review journal? (often includes “Rationale and design” in the paper title)  Mentioning the study protocol without a reference/citation/link is insufficient and should be “No” |
| **Standardised reporting guideline used** | Dropdown: Yes, No, Unsure | Does the paper refer to the use of CONSORT standardized reporting guideline? Variations of CONSORT or potentially the SPIRIT guidelines may also be mentioned. See the [EQUATOR network](https://www.equator-network.org/?post_type=eq_guidelines&eq_guidelines_study_design=experimental-studies&eq_guidelines_clinical_specialty=0&eq_guidelines_report_section=0&s=) if unsure. |
| **Ethical approval** | Dropdown: Yes, No, Unsure | Does the paper refer to obtaining research ethics or IRB (Institutional Review Board) approval?  An explicit statement of approval is required, and reference to conduct according to “ethical standards” is not sufficient. A statement of review or approval may not mention this relating to ethics/ethical, however this is acceptable if the indication is external oversight and approval of the study, which will likely incorporate ethical aspects of research. |
| **Funding source** | Dropdown:   - Government body/agency - University - Industry (pharmaceutical and device companies) - Charity - Multiple funders - Other - Explicitly report no funding - Not stated | What is the funding source reported?  An online search may be helpful to determine which category a reported funder fits into.  Use “Explicitly report no funding” for papers that include a funding statement, and report no specific funding. Use “Not stated” which papers that do not include information on funding.  Select Other if still unsure of which category and provide details in Funding comments. |
| **Funding comments** | Free text | Only need to fill in if Multiple funders or Other selected in previous column to provide further detail. |
| **Study characteristics & methods** |  |  |
| **Health condition/area (e.g. asthma, depression)** | Free text | Provide details of the health condition/topic that is the focus of the trial |
| **Health condition HRCS category if applicable** | Dropdown:   - Blood - Cancer and neoplasms - Cardiovascular - Congenital disorders - Ear - Eye - Infection - Inflammatory and immune system - Injuries and accidents - Mental health - Metabolic and Endocrine - Musculoskeletal - Neurological - Oral and Gastrointestinal - Renal and Urogenital - Reproductive health and childbirth - Respiratory - Skin - Stroke - Generic health relevance - Disputed Aetiology and Other | Categorise the response from the previous column, further details at:  <https://hrcsonline.net/health-categories/>  Select whichever you think fits best – we will do a final review of the actual free text from the previous column within each category at the end, so can pick up any issues. |
| **Clinical setting** | Dropdown:   - Primary care - Ambulatory (out-patient) - Hospital (in-patient) - Lab based - Multiple sites - Not stated - Other | In what setting was the study conducted?  This should be where the intervention is delivered, in the case of recruitment happening in a different setting e.g. an intervention associated with operative care should be in-patient.  If the majority of intervention delivery is in one setting (e.g. ambulatory care), with recruitment, baseline assessments and first doses occurring in a different setting, then it should be classified where the majority of intervention delivery occurs (with multiple sites only used where there is a more even split across two or more settings).  Can be inferred based on population/intervention using your clinical judgement e.g. intervention in neonates indicating hospital setting. |
| **RCT design** | Dropdown:   - Parallel - Cluster - Factorial - Crossover - Stepped wedge - Not stated - Other | What specific type of RCT design was used? Parallel design is the most common.  If multiple RCT designs in the same study, generally it would be appropriate to classify as the initial design where results are presented for this phase (e.g. a parallel two group study, where participants all crossover to just the treatment group at some point, could be classified as parallel.) |
| **Unit of randomisation** | Dropdown:   - Individual - Cluster/group - Body part - Not stated - Other | What was the unit of allocation i.e. was it randomizing patients, wards/clinics/GP practices, parts of the body (e.g. right eye and left eye) |
| **Blinding** | Dropdown:   - Open label - Single blind - Double blind | Whether the paper reports blinding of participants, care providers, and/or investigators. If no blinding is reported, select open label. If blinding of participants is reported, select single blind. If blinding of participants and care providers and/or investigators is reported, select double blind.  If multiple types of blinding at different phases, generally record the initial type of blinding e.g. a double blind study which becomes an open label study would be classified as double blind. |
| **Intervention type (see ISCRTN tab)** | Dropdown:   - Drug - Device - Procedure/Surgery - Behavioural - Genetic - Supplement - Biological/Vaccine - Service organisation/policy - Not stated - Other - Mixed | What category did the intervention of interest fall into?  Food or other dietary interventions can be included under supplement. |
| **Comparison group** | Dropdown:   - Alternative drug - Same drug, different dose/formulation etc - Placebo/sham - Alternative intervention - Usual care - No intervention - Multiple (no placebo) - Multiple (incl. placebo) - Not stated - Other | What category did the comparator(s) fall into?  In the following scenarios:   - Multiple arms of the same drug and different dose formulation, versus a placebo (e.g. Ramipril 10mg vs Ramipril 20mg vs placebo) 🡪 Categorise as placebo - Multiple comparator arms of different drugs, none of which are placebo (e.g. Ramipril 10mg vs Valsartan 80mg vs Olmesartan 10mg) 🡪 Multiple (no placebo) - Multiple comparator arms, one of which is placebo (e.g. Ramipril 10mg vs Valsartan 80mg vs Olmesartan 10mg) 🡪 Multiple (incl. placebo)   Same drug, different dose/formulation to be used for only cases where no alternative drug/treatment/placebo/no intervention/usual care arm included.  If they consider multiple drugs, at multiple doses, versus placebo, can disregard the multiple doses of the same drug, and consider this as Multiple (incl. placebo). |
| **Primary outcome** | Free text | A description of the primary outcome(s) e.g. HbA1c, mortality |
| **HRCS Research Activity Code** | Numerical value (e.g. 6.1) | What was the research activity aiming to do? This should be categorized per the HRCS activity classification (see tab or <https://hrcsonline.net/research-activities/>  For example a study evaluating the effect of a pharmaceutical would be coded as 6.1  For dietary interventions, 6.7 (evaluation of an intervention - complementary) is likely the most appropriate category.  For studies comparing two formulations of a drug, 6.1 (pharmaceuticals) is appropriate. |
| **Participants** |  |  |
| **Population** | Dropdown:   - Infants - Children - Adults - Older people - Pregnant women - Multiple - Not stated - Other | Who are the participants in the study? Where a population fits in more than category, please select the broader category. |
| **Total number randomised** | Numerical value | Report number of individuals randomized  If some randomized as protocol deviations and later withdrawn from the study, it is still appropriate to extract the actual number randomized. Take the value from the main body of the paper if there is an unexplained discrepancy with the abstract. |
| **Required sample size reached?** | Dropdown: Yes, No, Unsure, No sample size calculation | Did the total number randomized meet or exceed the calculated sample size? Normally there will be a sample size calculation presented in the Methods section.  Where conduct of a sample size is mentioned, but the calculated sample size is not reported, select “No sample size calculation” |
| **Number of Irish participants** | Numerical value | Number of participants from Ireland (if reported) |
| **Percentage of Irish participants** | Numerical value | Percentage of participants from Ireland (only record this if reported – no need to calculate based on number of participants) |
| **Total number of countries** | Numerical value | Number of countries where participants were recruited (if reported) |
| **Names of countries** | Free text | List names of all countries, separating each with a comma (if reported) |
| **Total number of centres** | Numerical value | Number of centres where participants were recruited (if reported) i.e. number of clinics/sites/hospitals. If the names of individual centres are listed out, please count overall number of centres, and Irish centres. |
| **Number of Irish centres** | Numerical value | Number of centres in Ireland (if reported)  If there is a discrepancy in number of (Irish) centres reported in the published paper compared to the registry, record the number reported in the paper, and make a note of the discrepancy in the queries column. Where only one source of this number is present, just record that number. |
| **Percentage of Irish centres** | Numerical value | Percentage of centres in Ireland (only record this if reported – no need to calculate based on number of centres) |
| **Total number of authors** | Numerical value | Total number of named authors on the paper (no need to count a group if included in the author list e.g. on behalf of the XXX investigators) |
| **Number of Irish authors** | Numerical value | Number of named authors with an affiliation within Ireland e.g. a university, hospital etc. |
| **Named investigators presented** | Dropdown: Yes, No, Unsure | Is there a list of names of all the investigators across sites, usually provided just before/after the References or as an appendix/supplementary material. |
| **Results** |  |  |
| **Baseline characteristics (Table 1) presented** | Dropdown: Yes, No, Unsure | Are characteristics of treatment groups at baseline provided? Typically this in the form of a table 1, listing each treatment group in a separate column. These details may be reported in a supplementary/appendix table, or simply in the text of the Results. |
| **Primary outcome positive or negative** | Dropdown: Positive, Negative, Unsure | Was a statistically significant effect on the primary outcome(s) reported?  In scenarios where no primary outcome specified, or multiple primary outcome specified, it is ok to make a judgement on whether largely positive or negative, or if very mixed, to select unsure, and note details in the other queries box. |
| **Corresponding Author** |  |  |
| **Name and affiliation** | Free text | Details of the corresponding author (name and institution) |
| **Email address** | Free text | Email address of the corresponding author |
| **Other queries** | Free text | Please detail any queries or uncertainties |

eTable 2. Study documentation and reporting and associated criteria

| **Variable** | **Criteria** |
| --- | --- |
| Ethical approval | Mention of the study having ethical approval was considered sufficient to show ethical approval. Reference to conduct according to “ethical standards” was not considered sufficient. |
| Trial registration number provided | Mention of any trial registration numbers (e.g. NCT, EUCTR, ISRCTN) was considered sufficient for this variable. |
| Protocol publication | If the relevant trial publication provided a link to a protocol or referenced a protocol published in a peer review journal, this was considered sufficient. |
| Use of a standardised reporting guideline | Mention of the use of the CONSORT reporting guideline or the ‘Standard Protocol Items: Recommendations for Interventional Trials’ (SPIRIT) reporting guideline was considered sufficient to show use of a standardised reporting guideline. |
| Sample size calculation | Consisted of two variables a) information provided and b) whether sample size was reached. |

eTable 3. Variables where it was not possible to make clear conclusions for two trials because the primary publication was not available

|  | Information not available | |
| --- | --- | --- |
| Variable | Trial A | Trial B |
| Named investigators presented | x |  |
| Baseline characteristics (Table 1) presented | x |  |
| Standardised reporting guideline used | x | x |
| Required sample size reached |  | x |
| Refers to published protocol |  | x |
| Ethical approval |  | x |
| Clinical trial registration number |  | x |

eTable 4. Type of comaparison groups used as part of trials

| Type of comparison group | % N |
| --- | --- |
| Placebo/sham | 25.1% (N=60) |
| Alternative drug | 20.9%, N=50 |
| Alternative intervention | 10.9%, N=26 |
| Usual care | 8.8% (N=21) |
| Same drug, different dose/formulation | 7.9% (N=19) |
| No intervention | 2.5% (N=6) |
| Multiple (two or more of groups above including placebo) | 11.3% (N=27) |
| Multiple (two or more of groups above but no placebo) | 12.6% (N=30) |

eBox 2. Source of the information on countries

For 96.7% (N=231) of trials the source of the information on countries was the primary paper, for 0.8% (N=2) it was the registry, and for 2.5% (N=6) of trials information on countries was gathered from both the registry and the paper

Information on the number of countries that participants were recruited from was provided in 99.6% (N=238) of trials, and 0.4% (N=1) of trials reported information that facilitated estimation of the number of countries.

eTable 5 Countries in which trials had at least one centre

| Country | Number of trials | % of all trials |
| --- | --- | --- |
| United Kingdom | 194 | 81.2% |
| Germany | 94 | 39.3% |
| Belgium | 85 | 35.6% |
| France | 84 | 35.1% |
| Spain | 79 | 33.1% |
| Netherlands | 74 | 31.0% |
| Italy | 70 | 29.3% |
| Canada | 65 | 27.2% |
| Australia | 60 | 25.1% |
| Sweden | 57 | 23.8% |
| United States | 57 | 23.8% |
| Poland | 56 | 23.4% |
| Austria | 45 | 18.8% |
| Denmark | 43 | 18.0% |
| Switzerland | 42 | 17.6% |
| Czechia | 39 | 16.3% |
| Hungary | 38 | 15.9% |
| Russia | 35 | 14.6% |
| Israel | 33 | 13.8% |
| Finland | 32 | 13.4% |
| South Africa | 32 | 13.4% |
| Norway | 31 | 13.0% |
| Argentina | 28 | 11.7% |
| Greece | 27 | 11.3% |
| New Zealand | 27 | 11.3% |
| Portugal | 23 | 9.6% |
| Romania | 23 | 9.6% |
| Brazil | 21 | 8.8% |
| Mexico | 19 | 7.9% |
| Turkey | 18 | 7.5% |
| Chile | 17 | 7.1% |
| Korea | 17 | 7.1% |
| Slovakia | 17 | 7.1% |
| Bulgaria | 14 | 5.9% |
| Taiwan | 14 | 5.9% |
| Japan | 13 | 5.4% |
| China | 12 | 5.0% |
| India | 12 | 5.0% |
| Slovenia | 12 | 5.0% |
| Ukraine | 12 | 5.0% |
| Colombia | 10 | 4.2% |
| Croatia | 10 | 4.2% |
| Hong Kong | 10 | 4.2% |
| Estonia | 9 | 3.8% |
| Singapore | 9 | 3.8% |
| Iceland | 9 | 3.8% |
| Malaysia | 8 | 3.3% |
| Lithuania | 7 | 2.9% |
| Peru | 6 | 2.5% |
| Serbia | 6 | 2.5% |
| Latvia | 6 | 2.5% |
| Thailand | 5 | 2.1% |
| Tunisia | 5 | 2.1% |
| Belarus | 5 | 2.1% |
| Luxembourg | 4 | 1.7% |
| Egypt | 3 | 1.3% |
| Philippines | 3 | 1.3% |
| Uruguay | 3 | 1.3% |
| Ecuador | 2 | 0.8% |
| Georgia | 2 | 0.8% |
| Lebanon | 2 | 0.8% |
| Nigeria | 2 | 0.8% |
| Saudi Arabia | 2 | 0.8% |
| Vietnam | 2 | 0.8% |
| Costa Rica | 2 | 0.8% |
| UAE | 2 | 0.8% |
| Cyprus | 2 | 0.8% |
| Ghana | 1 | 0.4% |
| Guadeloupe | 1 | 0.4% |
| Guatemala | 1 | 0.4% |
| Indonesia | 1 | 0.4% |
| Iran | 1 | 0.4% |
| Ivory Coast | 1 | 0.4% |
| Kenya | 1 | 0.4% |
| Moldova | 1 | 0.4% |
| Monaco | 1 | 0.4% |
| Pakistan | 1 | 0.4% |
| Panama | 1 | 0.4% |
| Paraguay | 1 | 0.4% |
| Qatar | 1 | 0.4% |
| Venezuela | 1 | 0.4% |
| Morocco | 1 | 0.4% |
| Bosnia and Herzegovina | 1 | 0.4% |
| Macedonia | 1 | 0.4% |
| Albania | 1 | 0.4% |
| Cuba | 1 | 0.4% |
| Sri Lanka | 1 | 0.4% |
| Uganda | 1 | 0.4% |
| Zimbabwe | 1 | 0.4% |
| Jordan | 1 | 0.4% |

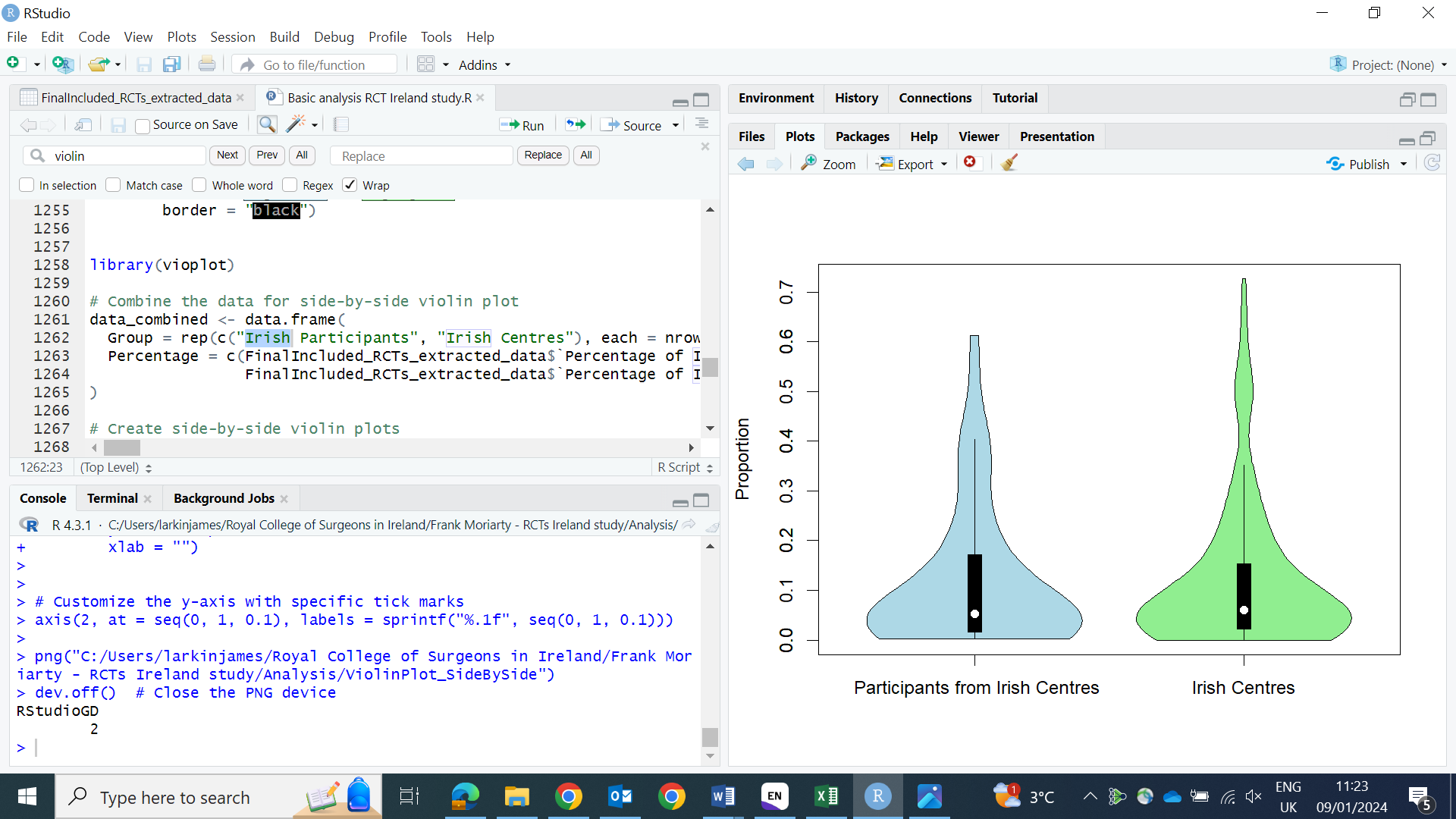

eFigure 1. Violin plots showing proportion of Irish centres and participants recruited from Irish centres in trials
